# Association Between Platelet Indices and Platelet Count in Patients with Immune Thrombocytopenia During Routine Follow-up

**DOI:** 10.64898/2026.02.04.26345597

**Authors:** Sufian J. Alalagy, Salem Altaeb

**Affiliations:** Department of Internal Medicine, Benghazi Medical Center, Benghazi, Libya

**Keywords:** Immune Thrombocytopenia, Platelet Count, Platelet Distribution Width, Mean Platelet Volume, Thrombocytopenia

## Abstract

**Aims:** To evaluate the association between platelet indices and platelet count severity in patients with primary immune thrombocytopenia during routine post-treatment follow-up.

**Methods:** This retrospective observational study included patients with primary immune thrombocytopenia followed at a single tertiary care center between 2011 and 2025. Demographic and laboratory data were obtained from medical records. Platelet count severity was categorized as less than 30 × 10^9/L, 30 to 100 × 10^9/L, and greater than 100 × 10^9/L. Platelet indices, including mean platelet volume (MPV) and platelet distribution width (PDW), were analyzed using the most recent complete blood count obtained during routine follow-up after treatment initiation. Continuous variables were summarized as median and interquartile range. Comparisons across platelet count categories were performed using the Kruskal-Wallis test with post hoc Mann-Whitney U testing. Correlation analysis and simple linear regression were also conducted.

**Results:** A total of 243 patients were identified, of whom 232 met the inclusion criteria. Platelet distribution width differed significantly across platelet count severity categories (Kruskal-Wallis p < 0.001) and demonstrated a strong inverse association with platelet count. Mean platelet volume also showed a statistically significant difference across platelet count groups (Kruskal-Wallis p = 0.007), although the association was weaker and less consistent compared with PDW. Regression analysis confirmed a significant association between platelet count and PDW.

**Conclusion:** Platelet distribution width is more closely associated with platelet count severity than mean platelet volume in patients with primary immune thrombocytopenia during routine post-treatment follow-up. PDW may represent a useful adjunctive laboratory parameter when interpreted alongside platelet count in routine clinical practice.

## Introduction

Immune thrombocytopenia (ITP) is an autoimmune disorder characterized by a persistently low platelet count, typically defined as <100 × 10□/L, and an associated but highly variable risk of bleeding [1]. It commonly manifests as isolated thrombocytopenia with purpura and, in some cases, hemorrhagic episodes. In routine clinical practice, the diagnosis is usually established by excluding other known causes of thrombocytopenia. ITP without an identifiable underlying condition is referred to as primary ITP, whereas secondary ITP occurs in association with external factors, including autoimmune diseases such as systemic lupus erythematosus, chronic infections such as HIV or hepatitis C virus infection and drug exposure [2]. Despite similar laboratory findings, the clinical course can vary considerably. Some patients tolerate very low platelet counts with minimal symptoms, while others experience bleeding at much higher levels, reinforcing the fact that platelet count alone does not fully capture disease behavior.

From a pathophysiological perspective, ITP is driven by increased peripheral platelet destruction and/or reduced or inadequate platelet production. Most patients are found to have autoantibodies directed against specific platelet membrane glycoproteins, leading to immune-mediated platelet clearance [3].

In recent years, attention has increasingly focused on platelet indices derived from routine complete blood counts (CBCs), particularly mean platelet volume (MPV) and platelet distribution width (PDW). Several studies have reported that patients with ITP have higher MPV and PDW values compared with healthy controls, suggesting the presence of larger and more heterogeneous platelets in this condition [4]. For example, Ntaios et al. found significantly elevated MPV and PDW in ITP patients compared with non-ITP thrombocytopenic patients [5]. However, the relationship between platelet indices and platelet count severity has not been consistent across studies and their clinical utility remains uncertain.

This uncertainty becomes especially apparent during routine clinical follow-up, where CBCs are obtained at variable time points and often after treatment initiation limiting longitudinal interpretation. Despite growing interest in platelet indices in immune thrombocytopenia, their interpretation in real-world clinical settings remains challenging. Therefore, the present study was designed to evaluate the association between platelet indices and platelet count severity in adult patients with immune thrombocytopenia during routine clinical follow-up.

## Methods

### Study Design and Setting

This was a retrospective observational study conducted at a single tertiary care center and included patients followed between January 2011 and December 2025.

### Study Population

Medical records of 243 adult patients evaluated for immune thrombocytopenia during routine clinical follow-up were reviewed.

### Inclusion and Exclusion Criteria

Adult patients (≥18 years) with a diagnosis of primary immune thrombocytopenia were eligible for inclusion. Patients with secondary causes of thrombocytopenia were excluded, including autoimmune diseases, hematologic malignancies, chronic infections, and other identifiable secondary causes. A total of 11 patients were excluded based on these criteria, resulting in 232 patients included in the final analysis.

### Data Collection

Demographic and laboratory data were obtained from medical records. Laboratory parameters derived from routine complete blood counts (CBC) included platelet count, mean platelet volume (MPV), platelet distribution width (PDW), hemoglobin level, and white blood cell count. All complete blood counts included in the analysis were obtained during routine post-treatment follow-up, and for patients with multiple results available, the most recent complete blood count was selected for analysis, reflecting routine clinical practice. Due to the retrospective nature of the study, MPV and/or PDW values were not available for all patients; analyses involving platelet indices were therefore conducted using available data only.

### Statistical Analysis

Statistical analyses were performed using IBM SPSS Statistics version 24. Continuous variables were assessed for normality using visual inspection and the Shapiro–Wilk test. As all continuous variables demonstrated non-normal distributions, they were summarized as median and interquartile range (IQR), while categorical variables were presented as frequencies and percentages.

Platelet count was analyzed both as a continuous variable and after stratification into clinically relevant categories (<30 × 10□/L, 30–100 × 10□/L, and >100 × 10□/L). Comparisons of platelet indices across platelet count categories were performed using the Kruskal–Wallis test. Where applicable, post-hoc pairwise comparisons were conducted using Mann–Whitney U tests with Bonferroni correction.

Associations between platelet count and platelet indices were evaluated using Spearman’s rank correlation coefficient. Simple linear regression analysis was subsequently performed to assess the relationship between platelet count and platelet distribution width (PDW), with PDW entered as the dependent variable and platelet count as the independent variable. Regression assumptions were assessed prior to analysis. All statistical tests were two-tailed, and a p-value <0.05 was considered statistically significant.

## RESULTS

The baseline demographic and clinical characteristics of the study cohort are summarized in Table 1.

**Table 1.** Baseline characteristics of patients.

| Characteristic | value |
| --- | --- |
| Number of patients | 232 |
| Age, median (IQR) | 39 (21) |
| Female sex, n(%) | 182 (78.4%) |
| Male sex, n(%) | 50 (21.6%) |
| Duration of ITP, median (IQR) | 7 (7) |
| Hemoglobin, g/dL, median (IQR) | 12.3 (2.5) |
| White blood cell count, $\times 10^9/L$, median (IQR) | 8.2 (5.5) |
| **Treatment during follow-up, n(%)** |  |
| No treatment | 39 (16.8%) |
| Steroids ± IVIG | 193 (83.2%) |

Platelet count severity was stratified into clinically relevant categories. Twenty-seven patients (11.6%) had platelet counts <30 ×10□/L, 75 patients (32.3%) had platelet counts between 30 and 100 ×10□/L, and 130 patients (56.0%) had platelet counts >100 ×10□/L. Mean platelet volume and platelet distribution width differed significantly across platelet count severity categories, with MPV showing a significant overall difference (Kruskal– Wallis p = 0.007) and PDW demonstrating a stronger association (Kruskal–Wallis p < 0.001), as summarized in Table 2.

**Table 2.** Platelet indices across platelet count severity groups.

| Platelet groups | MPV, median (IQR) | PDW, median (IQR) |
| --- | --- | --- |
| $<30 \times 10^9/L$ | 10.9 (2.8) | 18.1 (3.8) |
| $30\text{--}100 \times 10^9/L$ | 11.1 (2.2) | 17.4 (2.5) |
| $>100 \times 10^9/L$ | 10.1 (1.8) | 14.3 (4.3) |
| p-value | 0.007 | <0.001 |

Post-hoc pairwise comparisons with Bonferroni correction demonstrated a significant difference in mean platelet volume between patients with platelet counts of 30–100 ×10□/L and those with platelet counts >100 ×10□/L (p = 0.001), while no significant differences were observed between the <30 ×10□/L group and the other groups.

For platelet distribution width (PDW), post-hoc analysis revealed significantly higher values in patients with platelet counts <30 ×10□/L and 30–100 ×10□/L compared with those with platelet counts >100 ×10□/L (both p < 0.001), whereas no significant difference was observed between the <30 ×10□/L and 30–100 ×10□/L groups.

Spearman correlation analysis demonstrated a significant negative association between platelet count and platelet distribution width (r = −0.53, p < 0.001), as well as a weaker but significant negative association between platelet count and mean platelet volume (r = −0.25, p < 0.001). Analyses were performed using available data, resulting in varying sample sizes for MPV (n = 195) and PDW (n = 192).

In simple linear regression analysis, platelet count was significantly associated with platelet distribution width (unstandardized B = −0.011, p < 0.001), explaining approximately 13% of the variance in PDW (R^2^ = 0.13). Analysis was performed using available data (n = 192).

## Discussion

In this retrospective real-world study, we evaluated the association between platelet indices and platelet count severity in patients with primary immune thrombocytopenia during routine post-treatment follow-up. Our findings demonstrate that platelet distribution width (PDW) is strongly associated with platelet count severity, with significantly higher PDW values observed in patients with lower platelet counts. This association was consistent across group comparisons, correlation analysis and simple linear regression, supporting the concept that increased platelet size heterogeneity reflects enhanced peripheral platelet destruction and compensatory thrombopoiesis in ITP [6]. Mean platelet volume (MPV) also showed a statistically significant association with platelet count; however, this relationship was weaker and less consistent than that observed for PDW. This finding is in line with prior studies demonstrating the clinical and statistical significance of MPV in patients with thrombocytopenia, where MPV was shown to correlate with the underlying mechanism of platelet reduction [7]. Collectively, these results suggest that PDW may more reliably reflect platelet count severity than MPV in patients with ITP during routine clinical follow-up.

The strong inverse association observed between platelet count and PDW may be explained by the underlying pathophysiology of immune thrombocytopenia. PDW reflects the degree of platelet size heterogeneity and is considered a marker of anisocytosis within the circulating platelet population [8]. In ITP, immune-mediated peripheral platelet destruction stimulates compensatory thrombopoiesis, leading to the release of younger and larger platelets from the bone marrow alongside smaller, senescent platelets, thereby increasing platelet size variability and PDW values [9]. Several prior studies have reported higher PDW values in patients with immune thrombocytopenia and have demonstrated an inverse relationship between PDW and platelet count, supporting an association with platelet turnover and disease-related platelet heterogeneity rather than absolute platelet mass alone. For example, Kim et al. demonstrated that PDW increased as platelet counts decreased in immune thrombocytopenia [10]. Consistent with our results, Al-Sharifi et al. reported a significant inverse relationship between platelet count and PDW in a cohort of patients with thrombocytopenia, reinforcing the observation that increased platelet size heterogeneity is associated with more severe thrombocytopenia irrespective of underlying etiology [11].

In contrast to PDW, the association between mean platelet volume and platelet count severity in our study was statistically significant but comparatively weaker and less consistent. Although MPV differed across platelet count categories and showed a modest inverse correlation with platelet count, post-hoc analysis revealed significant differences only between selected platelet count groups, suggesting limited discriminatory capacity. Furthermore, Tang et al. reported that MPV, PDW, and other platelet indices reflect megakaryopoietic activity in the bone marrow of patients with ITP and may serve as markers of platelet production and destruction, although these indices alone have variable diagnostic performance [12]. A recent systematic review and meta-analysis by Walle et al. found that although mean platelet volume differed between patients with ITP and those with hypo-productive thrombocytopenia, the pooled sensitivity and specificity of MPV for differentiating ITP were modest, underscoring the limited and variable discriminatory value of MPV as a standalone marker in this context [13]. From a clinical perspective, our findings suggest that PDW may serve as a supportive parameter for contextualizing platelet count severity in patients with immune thrombocytopenia during routine follow-up. As PDW is automatically reported as part of the complete blood count, its interpretation does not require additional testing, cost or patient burden. In this context, Abdel Hi et al. demonstrated that PDW exhibited the highest diagnostic accuracy among platelet indices for distinguishing immune thrombocytopenia from other causes of thrombocytopenia, while MPV showed more limited performance, highlighting the relative robustness of PDW as a laboratory marker in thrombocytopenic states [14]. In real-world clinical practice, where platelet counts may fluctuate over time and are influenced by treatment exposure, PDW may therefore offer complementary insight into platelet heterogeneity and turnover when interpreted alongside absolute platelet counts rather than in isolation. However, consistent with observations by Khan et al., our findings do not support the use of PDW or MPV as standalone diagnostic or predictive markers. Instead, these indices should be regarded as adjunctive laboratory features that may contribute to a more nuanced hematologic assessment of patients with immune thrombocytopenia during routine clinical follow-up [15].

This study has several important strengths. The relatively large cohort of patients with primary immune thrombocytopenia enhances the robustness of the observed associations. In addition, the extended study period enabled evaluation of platelet indices across a broad spectrum of platelet counts encountered in routine clinical practice. The use of real-world post-treatment follow-up data further increases the clinical relevance of the findings to everyday hematologic care. Moreover, the consistency of results across multiple complementary analytical approaches, including group comparisons, correlation analysis and regression modeling, supports the internal validity of the observed association between platelet distribution width and platelet count severity.

Several limitations should be considered when interpreting these findings. The retrospective, single-center design limits generalizability and precludes causal inference. Platelet indices were assessed using the most recent complete blood count obtained during routine post-treatment follow-up rather than at diagnosis and may therefore have been influenced by treatment exposure and temporal variability. In addition, missing MPV and PDW values in a subset of patients reduced the sample size available for platelet index analyses. Finally, treatment-related factors were not examined in detail, and potential associations between platelet indices and specific therapies could not be evaluated.

## Conclusion

This study demonstrates that platelet distribution width is more closely associated with platelet count severity than mean platelet volume in patients with primary immune thrombocytopenia during routine post-treatment follow-up. While both platelet indices showed statistically significant associations with platelet count, PDW exhibited greater consistency across analytical approaches, suggesting that it may better reflect platelet heterogeneity and turnover in this setting. These findings support the interpretation of PDW as a useful adjunctive laboratory parameter when evaluating thrombocytopenia in routine clinical practice. Further prospective, multicenter studies are warranted to clarify the temporal behavior of platelet indices and to better define their role alongside established clinical and laboratory markers in immune thrombocytopenia.

## Data Availability

All data produced in the present work are contained in the manuscript.

## Acknowledgments

The authors declare no specific acknowledgments.

## Financial Disclosure

The authors received no financial support for the conduct or publication of this research.

## Notes

### Competing Interest Statement

The authors have declared no competing interest.

### Funding Statement

This study did not receive any funding.

### Author Declarations

This retrospective study was approved by the Research Ethics Board (REB), Benghazi Medical Center, Benghazi, Libya (Approval date: 19 November 2025; Approval number: NBC: 005.H.25.16).

